# Characterization of secretory phospholipase A2 inhibitory activity in *Tragia hispida* as potential therapeutic agent for treatment of dengue

**DOI:** 10.1101/2022.11.28.22282815

**Authors:** D. V. Dayangi Hemalika, U.G. Chandrika, Ajith M. Abeysekera, Sameera Samarakoon, Ananda Wijewickrama, Gathsaurie Neelika Malavige

## Abstract

**Background:** As secretory phospholipase A2 (sPLA2) was shown to be elevated in patients who progress to severe dengue, it would be important to evaluate the usefulness of therapeutics that inhibit sPLA2 enzymes to prevent progression to severe dengue.

**Objectives:** To explore the presence of sPLA2 inhibitors in plant extracts used in traditional medicine for treatment of fever in Sri Lanka.

**Study design:** Aqueous and butanol extracts of *Tragia hispida, Justicia adathoda* and tubers of *Cyperus rotundus* were screened for the presence of potential sPLA2 inhibitors using a commercial assay measuring sPLA2 activity.

**Results:** Both the aqueous (THA) and butanol extracts (THB) of *Tragia hispida* had sPLA2 inhibition levels comparable to the levels seen with the commercial sPLA2 inhibitor CAY10590. THB at concentrations of 0.1 µg/µL and 0.2 µg/µL, significantly reduced the sPLA2 activity (p<0.0001) in the sera of dengue patients and the inhibitory activity was significantly higher (p<0.0001) than of CAY10590. Thin layer chromatography of THB showed that it was likely to contain a mixture of flavonoid and phenolic compounds. HPLC displayed peaks at 3.207 min (λmax 222 nm, 272 nm) and 7.972 min (λmax 224 nm, 272 nm) were most likely to represent phenolics and that the peaks at 11.883 min (λmax 276 nm, 366 nm) and 16.898 min (λmax 254 nm, 370 nm) most likely to represent flavonoids.

**Conclusions:** *T. hispida* aqueous and butanol soluble fraction had potent sPLA2 inhibitory activities, which should be further explored for their potential to be used for treatment of dengue.

## Background

Dengue viral infections is one of the most rapidly emerging mosquitos borne infections which is estimated to infect 390 million individuals annually [1]. Although the age standardized infection rates, disability adjusted life years and mortality rates have increased over the last 30 years [2], there is no specific treatment for dengue. Due to climate change and rapid urbanization, the incidence of dengue is predicted to further increase in future [3]. Many dengue endemic countries experience seasonal outbreaks every year with the health care facilities becoming overwhelmed with the large number of dengue patients. Although most individuals infected with the dengue virus (DENV) develop mild illness, a significant proportion develop complications such as dengue haemorrhagic fever (DHF), organ dysfunction and bleeding [4]. However, as there are no prognostic markers to predict who is likely to develop severe disease and due to the non-availability of specific treatment, all dengue infected patients are serially monitored for early detection of complications for timely fluid management. Therefore, there is an urgent need to development of therapeutics for dengue.

Vascular leak is the hallmark of severe dengue, which leads to plasma leakage with fluid accumulation in pleural and peritoneal cavities, hypotension leading to shock and poor organ perfusion that contributes to organ dysfunction [5]. Endothelial dysfunction that leads to vascular leakage has shown to occur due to viral factors such as the secretory protein NS1 directly acting on the endothelial glycocalyx and activating immune cells to produce inflammatory mediators and inflammatory lipid mediators such as secretory phospholipase A2 (sPLA2s) and cytoplasmic phospholipase A2 enzymes [6-8]. In addition, a dysfunctional host innate immune response and pre-existing poorly neutralizing DENV specific antibodies also lead to endothelial dysfunction by acting on many different immune cells such as mast cells, monocytes and neutrophils. This in turn induces production of chymase, tryptase, leukotrienes, platelet activating factor (PAF) and sPLA2, which have shown to induce vascular leak [9-12]. There are several clinical trials that have been completed and several ongoing trials that have used repurposed drugs to inhibit PAF and those that stabilize mast cells [13, 14]. Apart from the above completed and ongoing trials, many patients in dengue endemic countries use herbal medicines to treat patients who present with fever.

*Tragia hispida*, which belongs to the plant family Euphorbiaceae, is a medicinal plant (known as “Welkahambiliya” in Sinhala) is used in the Sri Lankan traditional medicine to treat fever. Although the properties of *Tragia hispida* have not been characterized, a similar plant in the same family (*Tragia involucrate*) was shown to inhibit prostaglandin induced pain, have hepatoprotective effects against chemical induced hepatotoxicity in rats and antibacterial activity against many opportunistic bacteria [15]. *Justicia adathoda* is another plant used for treatment of fever and many inflammation related diseases in Sri Lanka and many other countries. This plant has shown to reduce carrageenan-induced inflammation in rats and had anti-pyrexic properties [16]. *Cyperus rotundus* too is included in many traditional medicines which is used to treat pain and many gynecological problems. This plant too has shown to have many anti-inflammatory and anti-pyretic properties [17]. However, the use and potential action of these plant extracts in treating patients with dengue has not been explored previously.

## Objectives

As many plant extracts are given in the aqueous form traditional medicine in Sri Lanka and many countries for treatment of fever, we sought to explore if any of these plant extracts were able to inhibit inflammatory mediators that associate with endothelial dysfunction in dengue. We previously showed that PAF was an important cause of vascular leakage and that phospholipase enzymes are important in the generation of PAF [18]. It was shown that the activity of the inflammatory lipid enzyme sPLA2 was significantly increased during the early illness in those who progressed to develop DHF [9, 10]. DENV NS1 protein was shown to induce sPLA2 activity and cPLA2 activity [6]. These phospholipase A2 enzymes hydrolyze membrane phospholipids thereby generating fatty acids, lysolipds and generate PAF. Therefore, we proceeded to explore if some of the plant extracts used in traditional medicine had any sPLA2 inhibitory activity.

## Study design

### Plant materials and preparation of aqueous, ethanol and n-butanol fractions

Fresh healthy aerial parts of *Tragia hispida, Justicia adathoda* and tubers of *Cyperus rotundus* were collected from Matara District, in the Southern Province of Sri Lanka during July to September 2017. *J. adathoda* and tubers of *C. rotundus* were authenticated by Nawinna Ayurveda research institute. The *T*.*hispida* plant was authenticated by the National Herbarium, Royal Botanical Garden, Peradeniya, Sri Lanka and a voucher specimen (Th/Matara/01), was deposited in the National Herbarium, Royal botanical garden, Peradeniya, Sri Lanka.

Fresh aerial parts (leaves and stems) of *T. hispida*, dried tubers of *Cyperus rotundus* and fresh aerial parts of *Justicia adathoda* were washed, air dried and subsequently oven dried at 40 ^0^C for 4 days and ground using a mechanical grinder. Two 300 g samples of *T. hispida* were extracted separately by refluxing the plant material for 2 hours in either 1.5 L of distilled water and 80 % ethanol to obtain an aqueous extract (THA) and an ethanol extract (THE) respectively. A portion of the THA extract was freeze dried and a portion of it was mixed with n-butanol to to obtain the butanol soluble fraction of the aqueous extract of *T. hispida* (THB).

In the preparation of *C. rotundus* plant extract, two 50 g samples of *C. rotundus* were extracted separately by refluxing the plant material for 2 hours in either 250 mL of distilled water and 80 % ethanol to obtain an aqueous extract (CRA) and an ethanol extract (CRE) respectively. CRE was evaporated under a vacuum to obtain a sticky brown semi-solid compound. The aqueous extract (CRA) was freeze-dried for 5 days to obtain a light brown coloured powder. *Justicia adathoda*, was prepared similarly with two 50 g samples extracted separately by 250 mL each of distilled water and 80 % ethanol by refluxing for 2 hours. Then 80 % ethanol extract (JAE) was evaporated under a vacuum to obtain brown semi-solid compound. The aqueous extract (JAA) was freeze-dried for 5 days to obtain a yellowish brown coloured powder.

### Assays for measuring secretory phospholipase A2 (sPLA2) activity and sPLA2 inhibitors

The sPLA2 activity was determined by a commercial assay (Abcam, UK), containing bee venom as the positive control (Catalog no: ab133089). The assay was carried out and interpreted according to the manufacturer’s instructions. All assays were carried out in triplicate. The sPLA2 activity was expressed as nmolmin^-1^mL^-1^. The sPLA2 inhibitory activity of plant extracts was compared with the commercially available sPLA2 inhibitor, CAY10590 (Cayman, USA).

### Characterization of Butanol fraction of *Tragia hispida* (THB) by High Performance Liquid Chromatography (HPLC)

HPLC was carried out on 1260 Agilent HPLC System, fitted with an Inert Sustain C18 (4.6 × 150 mm, 5 µm) column. In these assays, as the highest sPLA2 inhibitory activity was seen with THB, further characterization by HPLC was only carried out in this fraction. A solution of THB in methanol (400 ppm) was filtered first through a PTFE membrane filter (0.22µm) and then through a C18 cartridge filter (Bond Elut C18), and injected (10 µL) to the HPLC column. The elution was performed with 1% acetic acid and methanol at a ratio of 70:30 and the flow rate was maintained at 1mL/min throughout the procedure. The solvent composition was changed linearly after the 5 minutes to reach a composition of 1% acetic acid: methanol (30:70) at the end of 25 minutes. The chromatogram was monitored at 254 nm.

### Recruitment of patients with acute dengue infection

Blood samples were collected from 31 adult patients, with a duration of illness of ≤ 4 days from the National Institute of Infectious Diseases, Sri Lanka following informed written consent. All of them tested positive by the NS1 rapid antigen test (SD Diagnostics, South Korea). They were confirmed to have dengue by qRT-PCR as previously described and were found to be infected with dengue virus serotype 2 [19]. Serum was separated and stored at -20 ^0^C until further use. Ethical approval for the study was obtained from the Ethics Review Committee, Faculty of Medical Sciences, University of Sri Jayewardenepura (App. 12/15).

### Thin layer Chromatography (TLC) of THB

In the assays as the highest sPLA2 inhibitory activity was seen with THB, further characterization by TLC was only carried out on the THB fraction. THB was subject to thin layer chromatography on silica (Merck, EMD Millipore GF 254) using ethyl acetate: methanol: water (8: 1.2:1 v/v/v) as the developing solvent. The developed TLC was sprayed with Natural Products/ polyethylene glycol reagent as the visualizing agent.

### The Sulphorhodamine B assay (SRB assay) for assessing cytotoxicity

The cytotoxicity of THA was assessed using MRC-5 cell lines (ATCC, CCL-171, Virginia, USA) as previously described [20]. Briefly, MRC-5 cells were seeded at 5000 cells/well and co-cultured with varying concentrations of THA (25 to 400 µg/mL) diluted in DMSO. The cytotoxicity of THA was compared with that of Paclitaxel (doses ranging from 1.25 to 20 µg/mL), which was again diluted in DMSO. As DMSO itself it cytotoxic, the concentrations were kept below 0.05% in all experiments. Untreated MRC-5 cells were considered as the negative controls. All experiments were carried out in triplicate. The cytotoxicity was assessed by the Sulforhodamine B assay at 24, 48 and 72 hours according to the manufacturer’s instructions. The absorbance was measured at 540 nm using a microplate reader (Synergy TMHT multimode microplate reader, Bio Tek, USA). The percentage inhibition of cell growth for each concentration of the test substance was calculated. Half-maximal inhibitory concentrations (IC50) values were determined by analyzing sigmoid dose-response inhibition curves using GraphPad Prism software version 9.

### Statistical Analysis

GraphPad Prism version 9 was used for statistical analyses. As the data was not normally distributed non-parametric tests were used. In order to assess the differences in the inhibitory effects of the commercial sPLA2 inhibitor in comparison to different concentrations of THB, the Wilcoxon signed rank matched paired t test was used. P-values <0.05 were considered as statistically significant. The Holm-Sidak method, which corrects for multiple comparisons was used to compare the differences in the cell viability of MRC-5 cells treated with the plant extract compared to Paclitaxel.

## Results

### Screening of plant extracts for their potential sPLA2 inhibitory activity

We initially compared the sPLA2 inhibitory activities of the different extracts of *T. hispida* (THA, THE and THB), *C*.*rotundus* (CRA and CRE) and *J*.*adathoda* (JAA and JAE) with that of a commercial sPLA2 inhibitor (CAY 10590) on bee venom. Bee venom is known to have high levels of sPLA2 IIA. The inhibitory activity of each extract at different concentrations on bee venom sPLA2 is shown in table 1. We found that the extracts of *T. hispida* (THA and THE), showed the highest sPLA2 activity, which was comparable to the inhibition levels of CAY 10590. Of the *T. hispida extracts*, THB showed the highest level of inhibition of the sPLA2 activity and therefore, we proceeded with further analysis of this extract.

**Table 1:** The sPLA2 inhibitory activity of different fractions of *T. hispidia, C. rotundus* and *J. adathoda*. The sPLA2 inhibitor activity was assessed in *Tragia hispida*-80% ethanol extract (THE), aqueous extract (THA), *Cyperus rotundus* - 80% ethanol extract (CRE), aqueous extract (CRA), *Justicia adatoda* - 80% ethanol extract (JAE), aqueous extract (JAA). All extracts were assayed in two different concentrations, 0.1 µg/µL and 0.2 µg/µL in triplicates.

| **Name of plant extract**<br>± SD | Inhibitor concentration<br>0.1/ $\mu\text{g}/\mu\text{L}$ (C1)<br>sPLA2 activity / $\mu\text{mol}/\text{min}/\text{mL}$ | Percentage of inhibition of sPLA2 activity of bee venom at C1 concentration | Inhibitor concentration<br>0.2/ $\mu\text{g}/\mu\text{L}$ (C2)<br>sPLA2 activity / $\mu\text{mol}/\text{min}/\text{mL}$ | Percentage of inhibition of sPLA2 activity of bee venom at C2 concentration |
| --- | --- | --- | --- | --- |
| THA | $10.41 \pm 4.6$ | 54% | $8.83 \pm 4.9$ | 61% |
| THB | $9.22 \pm 8.5$ | 59.3 % | $3.41 \pm 1.9$ | 84.9 % |
| THE | $12.60 \pm 8.9$ | 44.3 % | $9.83 \pm 12.9$ | 56.5 % |
| CRA | 20.10 ± 9.7 | 11.2 % | 18.71 ± 8.1 | 17.3 % |
| CRE | 19.17 ± 8.1 | 15.3 % | 17.53 ± 13.3 | 22.6 % |
| JAA | 20.27 ± 18.7 | 10.4 % | 17.39 ± 11.1 | 23.2 % |
| JAE | 19.07 ± 3.34 | 15.8 % | 17.89 ± 2.6 | 21 % |
| CAY10590<br>(commercial<br>inhibitor) | 9.45 ± 8.4 | 58.2 % | * | * |

### Inhibitory effect of THB fraction on sPLA2 activity of serum from dengue patients

We previously showed that patients with dengue had high levels of sPLA2 activity during the febrile phase of dengue infection and therefore, serum samples were obtained from patients (n=31) with a duration of symptoms of ≤ 4 days [9]. The inhibitory effect of THB was assessed at concentrations of 0.1 µg/µL and 0.2 µg/µL on dengue patient sera. Both concentrations of THB significantly reduced the sPLA2 acitivity (p<0.0001) in the sera of dengue patients (Figure 1). The inhibition of sPLA2 acitivity of both concentrations of THB was significantly lower (p<0.0001) than the inhibition observed with the commercial sPLA2 inhibitor, CAY10590 (Figure 1).

**Figure 1:**
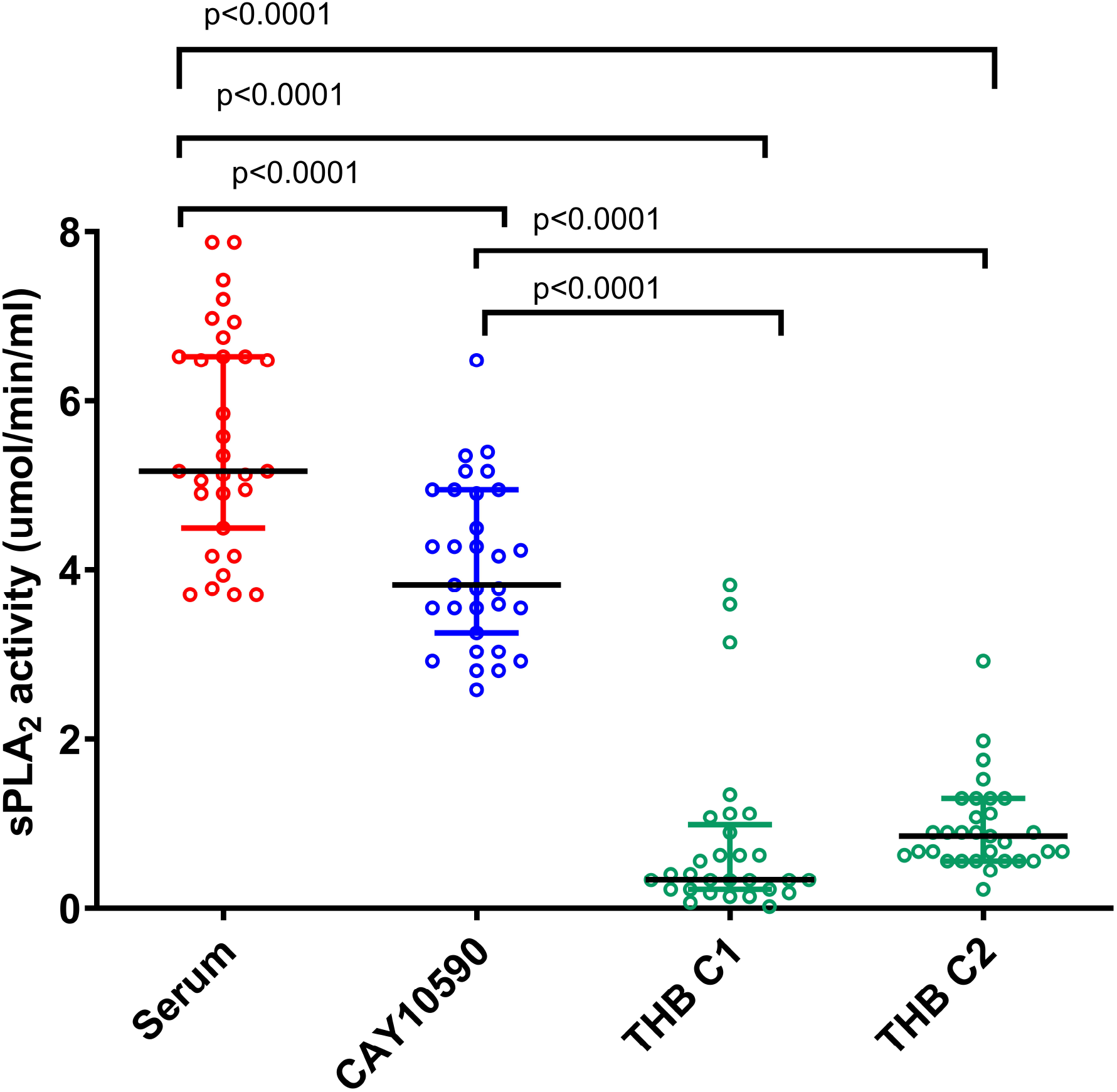
sPLA2 inhibitory activity of the butanol soluble fraction of the aqueous extract of *T. hispida* (THB) in patients with acute dengue infection. The sPLA2 inhibitory activity was measured in patients with acute dengue (n=31), and the inhibitory activity of THB at a concentration of 0.1 µg/µL (THB C1) and 0.2 µg/µL (THB C2) was compared with a commercial sPLA2 inhibitor (CAY10590). The error bars represent the median and the inter quartile range.

### Characterization secondary metabolites of butanol fraction of *Tragia hispida* (THB)

Although there chemical composition of *T. hispida*, is not known it has been shown that related species such as *T. involucrata* is rich in flavonoids, particularly in several glycosides and derivatives of quercetin [15, 21]. Flavonoids have shown to inhibit sPLA2 activity, and has shown to inhibit sPLA2 in synovial fluid but not from pancreatic fluid, showing that *T*.*hispidia* is most likely to have compounds that inhibit sPLA2 II group of enzymes [22]. The chemical composition of *T. hispida* (THB fraction) was characterised using thin layer chromotagraphy (TLC), which showed several poorly resolved yellow, orange and blue bands, indicating that it was likely to contact a mixture of flavonoid and phenolic compounds.

Since it was difficult to identify the possible compounds based on the TLC profile alone, HPLC was carried out to confirm the presence of those secondary metabolites such as flavinoids and phenolic compounds. The diode array UV showed the spectra of the four major peaks in HPLC profile. The peaks at 3.207 min (λmax 222 nm, 272 nm) and 7.972 min (λmax 224 nm, 272 nm) were most likely to represent phenolics and that the peaks at 11.883 min (λmax 276 nm, 366 nm) and 16.898 min (λmax 254 nm, 370 nm) most likely to represent flavonoids.

### SRB assay for cytotoxicity of the aqueous extract of *Tragia hispida* (THA)

Although the THB fraction gave the highest sPLA2 inhibitory activity, we sought to test the cytotoxicity of the aqueous extract of *T. hispida* (THA), as this plant is prepared as an aqueous extract in traditional medicine in Sri Lanka and THA is most likely to represent what is consumed in practice. The cytotoxicity was assessed using the SRB assay, comparing the cytotoxicity with that of Paclitaxel. THA was used at concentrations of 25, 50, 100, 200 and 400 μg/mL while Paclitaxel was used as a concentration of 1.25, 2.5, 5, 10, 20 μg /mL. The viability of MRC-5 cells at different concentrations at different time points with THA and Paclitaxel is shown in figure 4. The differences in the viability of the MRC-5 with the different concentrations of the drugs at different time points was assessed using the Holm-Sidak method which corrects for multiple comparisons which showed that the cell viability was significantly higher at 72 hours in those treated with THA compared to those treated with Paclitaxel (p=0.0005). The IC_50_ values for inhibition of cell growth, at 24, 48 and 72 hours were assessed and are shown in table 2.

**Table 2:** The IC_50_ values of THA on MRC-5 cell lines compared to Paclitaxel. The MRC-5 cell lines were treated with THA (25, 50, 100, 200 and 400 μg/mL) and Paclitaxel (1.25, 2.5, 5, 10, 20 μg /mL) and the cell proliferation was measured at 24, 48 and 72 hours by the SRB assay. All experiments were done in triplicate.

| Test Substance | IC<sub>50</sub> at 24h<br>/µgmL<sup>-1</sup><br>Mean (±SD) | IC<sub>50</sub> at 48h<br>/ µgmL<sup>-1</sup><br>Mean (±SD) | IC<sub>50</sub> at 72h<br>/ µgmL<sup>-1</sup><br>Mean (±SD) |
| --- | --- | --- | --- |
| THA | 4400 ± 0.4 | 445± 0.06 | 199 ± 0.03 |
| Paclitaxel (positive<br>control) | 217 ± 0.3 | 3.8 ± 0.1 | 0.0084 ± 0.5 |

**Figure 2:**
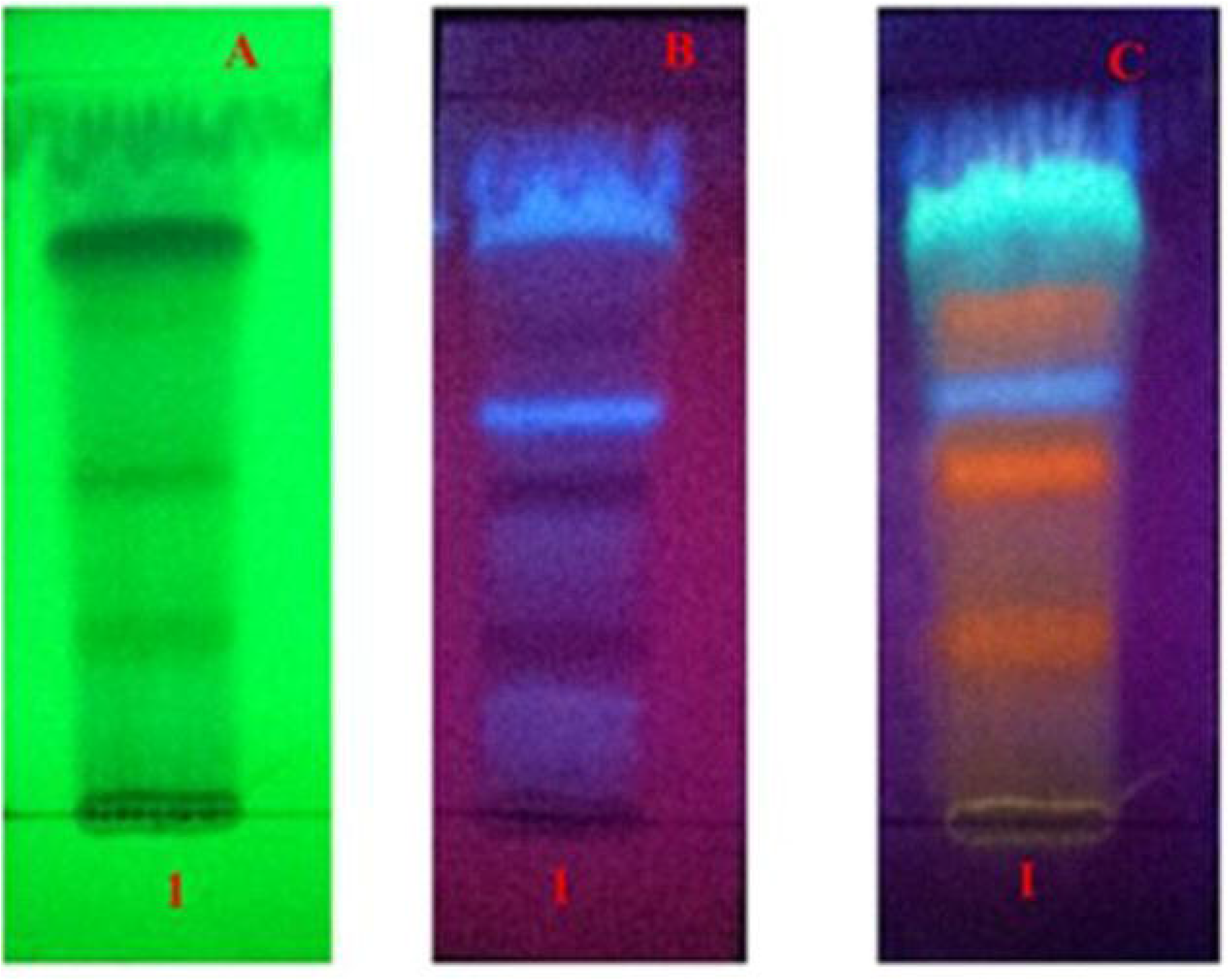
Thin layer chromatography profile of the butanol soluble fraction of the aqueous extract of *T. hispida* (THB) The TLC profile of THB was developed under the solvent system, ethyl acetate: methanol: water (8: 1.2:1 v/v/v). It was visualized as A-under 254 nm UV light, B-under 365 nm UV light and C-under 365 nm UV light after spraying with NPR/PEG reagent.

**Figure 3:**
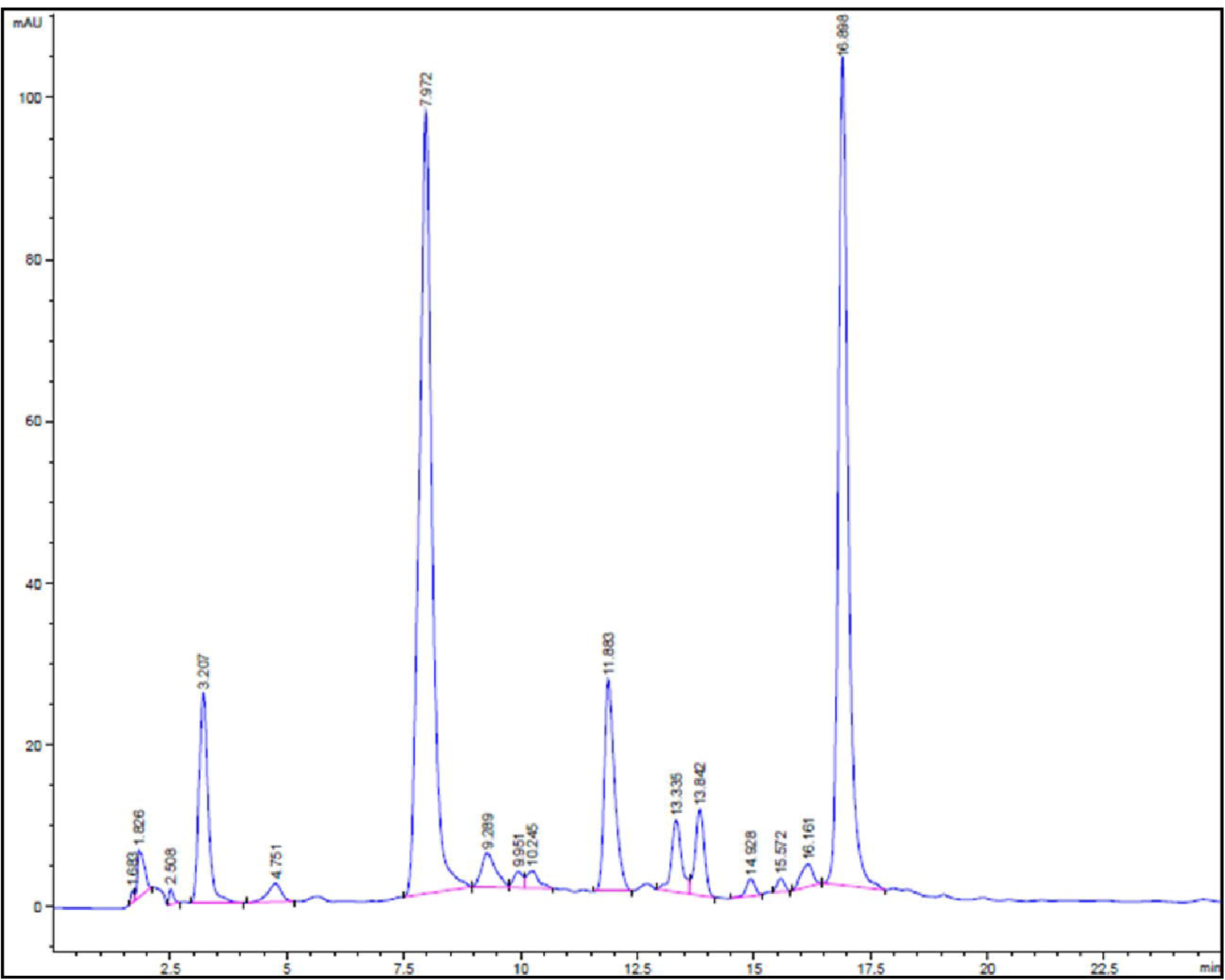
The HPLC chromatogram of butanol soluble fraction of the aqueous extract of T. hispida (THB) The HPLC chromatogram of butanol soluble fraction of the aqueous extract of *T. hispida* (THB) was obtained along with the diode array UV spectra of the four major peaks, characterized as phenolics and flavonoids.

**Figure 4:**
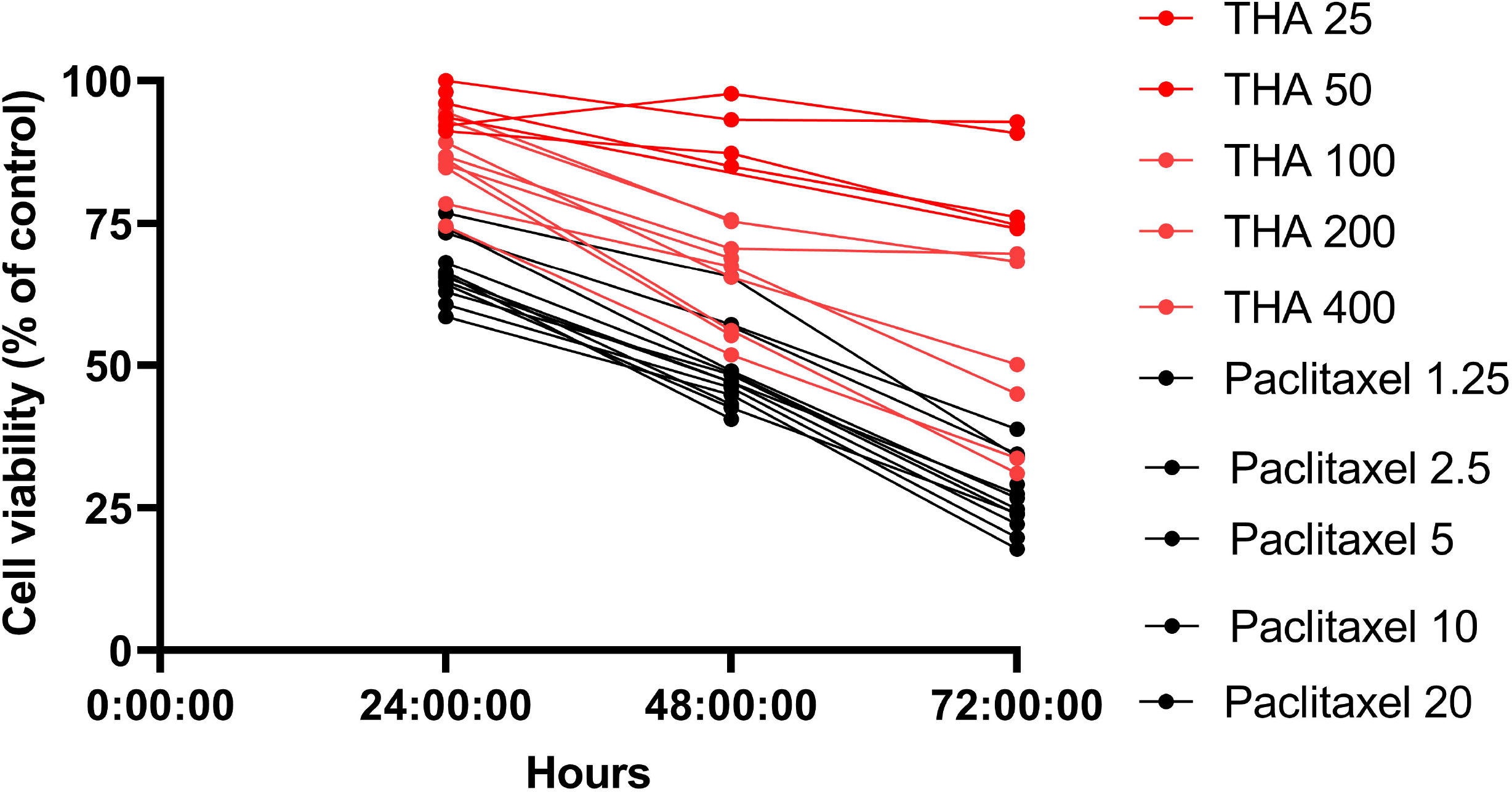
The SRB assay on MRC-5 cell lines for evaluation of toxicity of THA in comparison to Paclitaxel. The viability of the MRC-5 cell lines treated with varying concentrations of THA and Paclitaxel was assessed at 24, 48 and 72 hours from treatment. The different concentrations at different time points were carried out in triplicate.

## Discussion

In this study we investigated the potential sPLA2 inhibitory activity of several plant extracts that are used in traditional medicine in Sri Lanka and found that *T. hispida* aqueous and butanol soluble fraction had potent sPLA2 inhibitory activities. In fact, it had significantly higher sPLA2 inhibitory activity than the commercial sPLA2 inhibitor CAY10590, when assessed in dengue patient sera. The HPLC analysis showed that *T. hispida* butanol fraction contained many flavonoid compounds, which have previously shown to inhibit sPLA2 activity [22]. Snake venom is known to be a rich source of sPLA2 enzymes and aqueous solution of leaves and roots of *Tragia involucrata* have been taken orally by certain tribes in Tamil Nadu to treat snake bites [23]. However, the presence of sPLA2 inhibitory activity of these plant extracts had not been characterized previously.

The phospholipase A2 enzymes have many inflammatory actions in addition to generation of PAF, which has shown to be an important mediator of endothelial dysfunction in dengue [10]. sPLA2 is an acute phase protein, with a wide range of inflammatory effects [18]. Lipopolysaccharide (LPS), hypoxia and cytokines have shown to induce its activity [24, 25]. We have previously shown that the sPLA2 activity is significantly higher during early illness (72 to 84 hours since onset of illness) in patients who subsequently proceed to develop DHF [9]. In addition to its widely known inflammatory effects, sPLA2 has also shown to activate cytoplasmic PLA2 in mast cells, which in turn could contribute to generation of PAF and other arachidonic acid metabolites such as leukotrienes [26]. PAF acted synergistically with sPLA2 to induce neutrophil exocytosis, thereby is likely to further contribute to endothelial dysfunction and elevated cytokines seen in dengue [27]. Leukotrienes (LTE4) levels were shown to be higher in patients who proceeded to develop DHF, and the LTE4 levels continued to rise in patients with DHF[19]. Since sPLA2 appears to have a wide range of activities in the pathogenesis of endothelial dysfunction and severe dengue, drugs which inhibit sPLA2 could be of potential therapeutic value.

In this study we have shown that the leave extract of *T. hispida* which has been used for centuries to treat fever and inflammatory diseases in traditional medicine has potent sPLA2 inhibitory activity and in the cell cytotoxicity assays, it was shown to have minimum toxicity for mammalian cell lines. However, we could not isolate the exact compound in the extract of *T. hispida* that had these functions. Since the whole leave extract appears to be safe due to long term consumption and based on the evidence of in vitro studies, it would be important to carry out double-blind placebo controlled clinical trials to evaluate its efficacy in treating patients with acute dengue. In addition, it would be most important to also identify the exact chemical structure of this compound that had sPLA2 inhibitory activity.

## Data Availability

All data produced in the present work are contained in the manuscript

## Acknowledgements

We are grateful to the Centre for Dengue Research, University of Sri Jayewardenepura and University Grant Commission (grant number: UGC/DRIC/PG/2015(i)/OUSL/01) for funding this study. The authors thank S. B. R. C. Samarakoon of the Research and Development laboratory of Link Natural Products Ltd. for his assistance in developing the HPLC fingerprint for THB.

